# Prevalence and factors associated with use of prophylaxis for HIV and sexually transmitted infections among sexual health service attendees in England: findings from a cross-sectional observational study

**DOI:** 10.1101/2025.07.31.25332508

**Authors:** James Bell, Manik Kohli, Caisey Pulford, Dana Ogaz, Elizabeth Williams, Vanessa Apea, Nelson David, Emmi Suonpera, Richard Gilson, Catherine H. Mercer, Gwenda Hughes, David Dunn, Ed Showler, Hamish Mohammed, John Saunders

## Abstract

**Introduction:** HIV pre-exposure prophylaxis (PrEP) demonstrates the value of biomedical interventions as part of combination prevention. Some gay, bisexual and other men who have sex with men (GBMSM) use antibiotics to prevent sexually transmitted infections (STIs), primarily as post-exposure prophylaxis (PEP). We aimed to understand variations in awareness and use of HIV PrEP and STI PEP/PrEP among people attending specialist sexual health services (SHS) in England.

**Methods:** We collected data from SHS attendees aged >16 years in London and the East of England using an online questionnaire between April 2022 and December 2023, including awareness and use of HIV-PrEP and STI PEP/PrEP. We ran regression analyses: one to identify demographic factors associated with STI PEP/PrEP use among all SHS attendees, and a second to identify behavioural factors (controlling for socio-demographic factors) associated with antibiotic prophylaxis use among GBMSM.

**Results:** The study recruited 1,732 participants (50% GBMSM, 12% men who had reported sex with women only [MSW], 31% women). 74% had heard of HIV-PrEP but this varied by group (GBMSM:98%, MSW:39%, women:49%). 34% of GBMSM had heard of STI PEP/PrEP and 11% reported using it (MSW:15%/5%, women:18%/4%). 61% of STI PEP/PrEP users reported taking doxycycline, mostly to prevent chlamydia, gonorrhoea and syphilis. 80% of all participants reported being somewhat/very likely to use event-based STI PEP/PrEP, with a higher proportion of GBMSM saying they would be likely to use STI PEP/PrEP in this way. In the behavioural regression model of GBMSM, use was independently associated with reporting higher-risk sexual behaviour.

**Conclusion:** We found that a sizeable minority of GBMSM already use STI PEP/PrEP but some MSW and women also use or have an interest in antibiotic prophylaxis, suggesting potential benefits may exist for populations other than GBMSM. Communication should therefore should emphasise appropriate antibiotic use and the risks of antibiotic resistance among all populations.

**Key messages:** *KWhat is already known on this topic:*

- Despite widespread availability of HIV-PrEP, heterosexual men and women with a need for HIV-PrEP are less likely to have this met than GBMSM
- Some GBMSM use antibiotics to prevent bacterial STI infections in the absence of guidelines on STI PEP/PrEP in England

*What this study adds:*

- Awareness of HIV-PrEP is low among heterosexual men and women
- The study shows that some heterosexual men and women are currently using or have an interest in STI PEP/PrEP

*How this study might affect research, practice or policy:*

- This study could inform HIV-PrEP awareness activities for people at risk of HIV
- It could also inform communication strategies for any future introduction of STI PEP/PrEP

## Introduction

The use of biomedical interventions to prevent HIV and sexually transmitted infections (STIs) is becoming more widespread in England. In 2023 96,563 people initiated or continued HIV pre-exposure prophylaxis (HIV-PrEP) use (1). Despite increasing uptake, there is evidence that access to HIV-PrEP is not equitable: heterosexual men and women are less likely to have their need for HIV-PrEP identified during a visit to a sexual health service (SHS) than gay, bisexual and other men who have sex with men (GBMSM) (2).

STI prophylaxis is the use of antibiotics as post-exposure prophylaxis (PEP) or pre-exposure prophylaxis (PrEP) to reduce the risk of acquiring certain bacterial STIs. Three trials of doxycycline post-exposure prophylaxis (200mg administered within 72 hours of condomless sexual contact) in GBMSM and transgender women demonstrated efficacy in preventing chlamydia, gonorrhoea and syphilis. A single study among cisgender women did not result in decreased STI diagnoses, although this finding may have been due to suboptimal adherence in the intervention arm (3-6). Amid increasing interest in STI PEP and PrEP several countries now have guidelines or position statements supporting the use of doxycycline PEP for some individuals (7-12).

Although use of antibiotics to prevent syphilis has only recently been endorsed by guidelines in England, surveys from 2018-2021 suggest that between 4% and 9% of GBMSM or HIV-PrEP users in England had ever used antibiotic STI prophylaxis, and over 20% may be using antibiotics this way in other countries (13-19). Current prevalence estimates are based on samples of GBMSM, HIV-PrEP users or users of sexual networking apps, meaning there are no published studies from the UK that have specifically explored antibiotic use to prevent STIs in cisgender heterosexual men and women. There is also an incomplete recognition of the factors associated with the use of antibiotics to prevent STI as much of the existing evidence is based on surveys with small sample sizes.

The primary aim of this study was to estimate the prevalence of antibiotic STI prophylaxis awareness and use among SHS users, disaggregated by gender and sexual orientation. The study also aimed to explore the sociodemographic and behavioural factors associated with antibiotic use for STI prevention, explore attitudes towards antibiotic prophylaxis use and describe awareness and use of HV-PrEP.

## Methods

### Data collection

Data were collected using a cross-sectional self-administered anonymous online survey among SHS attendees at 19 SHS across London and the East of England. These services were selected because a large proportion of the GBMSM population in England access these services, and previous surveys have shown that people are using antibiotic STI prophylaxis in these areas (13-17). Service users who were aged ≥16 years and who were able to read English were eligible to take part. Attendees were invited to complete the survey after a telephone or face-to-face sexual health consultation. They were sent a text message containing a link to the survey or could scan a QR code in the clinic on their device. Posters advertising the study were placed in clinics, and researchers working at the SHS also approached participants directly during their visits.

The primary aim of the study was to estimate prevalence of STI prophylaxis use among SHS attendees, so a formal power calculation was not applicable. Using an assumed 8% prevalence among GBMSM (13-17), 1,250 responses would provide approximately 100 STI prophylaxis users among respondents. No published data existed at the time of the survey on STI prophylaxis use among other cisgender heterosexual men and women, so we sought to collect at least 100 responses from each group.

The survey contained questions on the sociodemographic characteristics of participants, their use of STI and HIV prophylaxis, their HIV and STI testing behaviour, sexual relationships and partnerships, and recreational drug use. The research question and items within the survey relating to willingness to use antibiotics to prevent STIs were developed with members of key communities at risk of STIs and HIV as well as members of community based organisations representing these communities. These questions have also been used in online, community based, cross sectional surveys delivered by UKHSA in partnership with HIV Scotland, PrEPster and iwantPrEPnow and the NIHR BBSTI Health Protection Research Unit

### Statistical analysis

Statistical analysis was carried out in Stata v18 (StataCorp LLC, College Station, TX, USA). In cases of incomplete responses to the survey, records were retained if the participant had answered the questions about STI antibiotic prophylaxis knowledge and usage (meaning that item non-response varies by question).

Participants were categorised into groups based on their self-described gender and sexual orientation: gay, bisexual and other men who have sex with men (GBMSM), men who have sex with only women (MSW), women, and those who describe themselves in another way or did not wish to provide a response. Participants were considered transgender if their gender identity was different than their sex assigned at birth. The demographic characteristics of the study sample were compared with the cohort of people who visited the clinics during the recruitment period to understand if the sample was representative of all clinic attendees. These data were extracted from the GUMCAD surveillance system.

We described awareness and use of HIV-PrEP and antibiotics for STI prophylaxis, including how these varied over the period of recruitment. For the subset of participants who had ever used antibiotics for STI prevention, we reported which antibiotics were used, which dosing schedule was adopted, the source of the antibiotics, if a healthcare provider was consulted before use, the timing of first and last doses, and which STIs the participant was trying to prevent.

We coded the results of an open-text question about the reasons for using antibiotics as STI prophylaxis, using themes identified inductively. Responses mentioned by fewer than 5% of participants were not reported. We then reported the likelihood and the level of concern with taking daily or event based antibiotic prophylaxis among the whole study sample. The above analyses were disaggregated primarily by gender and sexual orientation category (GBMSM, MSW and women), but differences by HIV status, HIV PrEP use, age, ethnicity, clinic location, and STI diagnoses in the last year (as a proxy for high sexual risk) were reported if noteworthy. Tables with full results are given in the Supplementary Materials.

An examination of the responses to the open-ended question on why participants had decided to use STI PEP/PrEP revealed potential misunderstanding among some participants. A sensitivity analysis was conducted to re-estimate the percentage of participants using STI-PEP/PrEP by excluding responses which referred only to emergency contraception, urinary tract infections, HIV-PrEP, STI treatment and antibiotic use as a result of partner notification. Responses were screened by JB and CP and discrepancies were resolved through discussion.

Two regression analyses were done. The first regression aimed to understand demographic factors associated with antibiotic prophylaxis use. We conducted bivariate and multivariable logistic regressions with a dichotomous variable indicating whether the participant had ever used antibiotic STI prophylaxis as the outcome. Age group (with age 25-34 as the reference group because it contains the highest number of participants), gender and sexual orientation category, ethnicity, if participants were born in the UK, location of clinic visited, years of education and employment status were considered explanatory variables.

A second logistic regression analysis was done to understand behavioural factors associated with antibiotic prophylaxis use among GBMSM with the same outcome variable as the first analysis. This analysis was not repeated for the other groups due to small sample sizes. In the bivariate models we considered HIV PrEP use and HIV status, STI diagnosis in the last year, number of partners in the last three months, drug use during sex and injectable drug use as explanatory variables. In the fully adjusted model, we included HIV status and HIV PrEP use, and created a composite variable for STI risk to avoid multicollinearity between predictors. A participant was considered to be at high risk if they reported at least two of the following: were diagnosed with an STI in the last year, had 11 or more partners in the last 3 months (as a proxy for high numbers of sexual partners), or reported sexualised drug use or injectable drug use (20, 21). Reporting of anal or vaginal sex without a condom was excluded due to the high number of GBMSM participants reporting this behaviour. The multivariable model also controlled for age, ethnicity and clinic location, as these variables may confound the association of interest. For all regression analyses we reported odds ratios and 95% confidence intervals.

### Ethics

Participants were given information about the study at the beginning of the survey link and asked to tick a box confirming their agreement to take part. Completion and submission of the survey denoted implied consent. No personal identifiers such as name or full address were collected in the survey. The study was approved by the NHS Research Ethics Committee (ref: S-1665).

## Results

### Sample Characteristics

Between April 2022 and December 2023 1,732 participants were recruited into the study. 50% described themselves as GBMSM, 12% as MSW, 31% as WSM and 0.4% as WSW (Table 1). The remaining 7% described themselves in another way or did not provide a response. The majority (64%) of participants were aged 25-44, were White (71%), were born in the UK (58%), attended a clinic in London (82%), had more than 2 years of post-16 education (75%) and were employed (81%). In total, 12% of participants were living with HIV (21% of GBMSM), 37% had received an STI diagnosis in the last year (50% of GBMSM) and 16% had had eleven or more sexual partners in the last 3 months (27% of GBMSM). Most (82%) reported anal or vaginal sex without a condom in the last three months, 34% had used drugs in the last year and 4% had ever used injectable drugs (49% and 6% respectively among GBMSM).

**Table 1:**
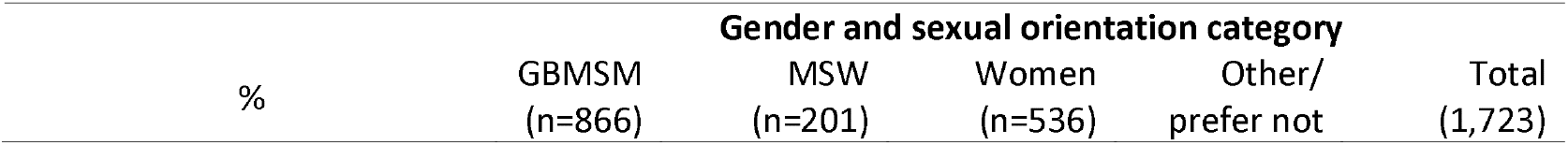
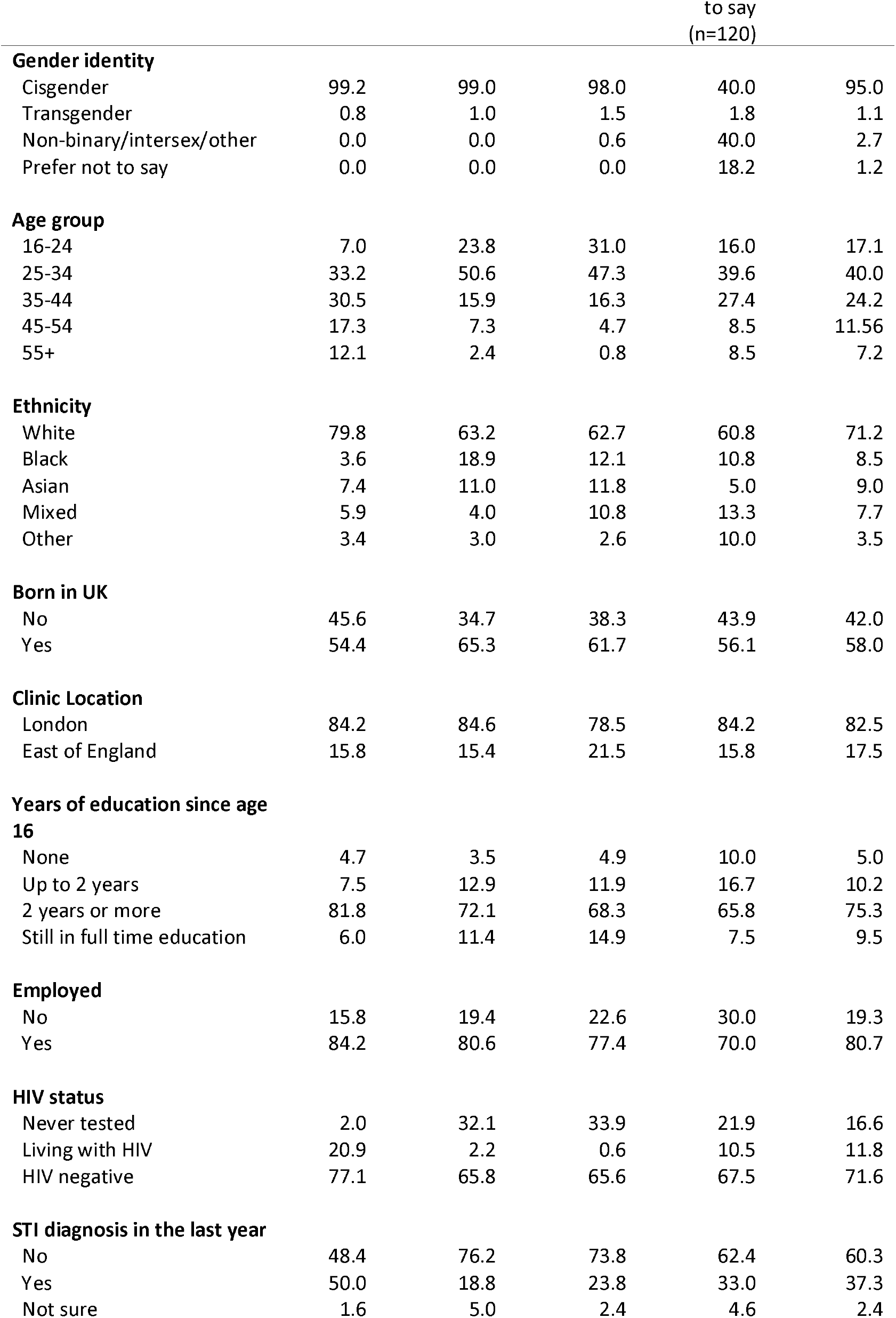
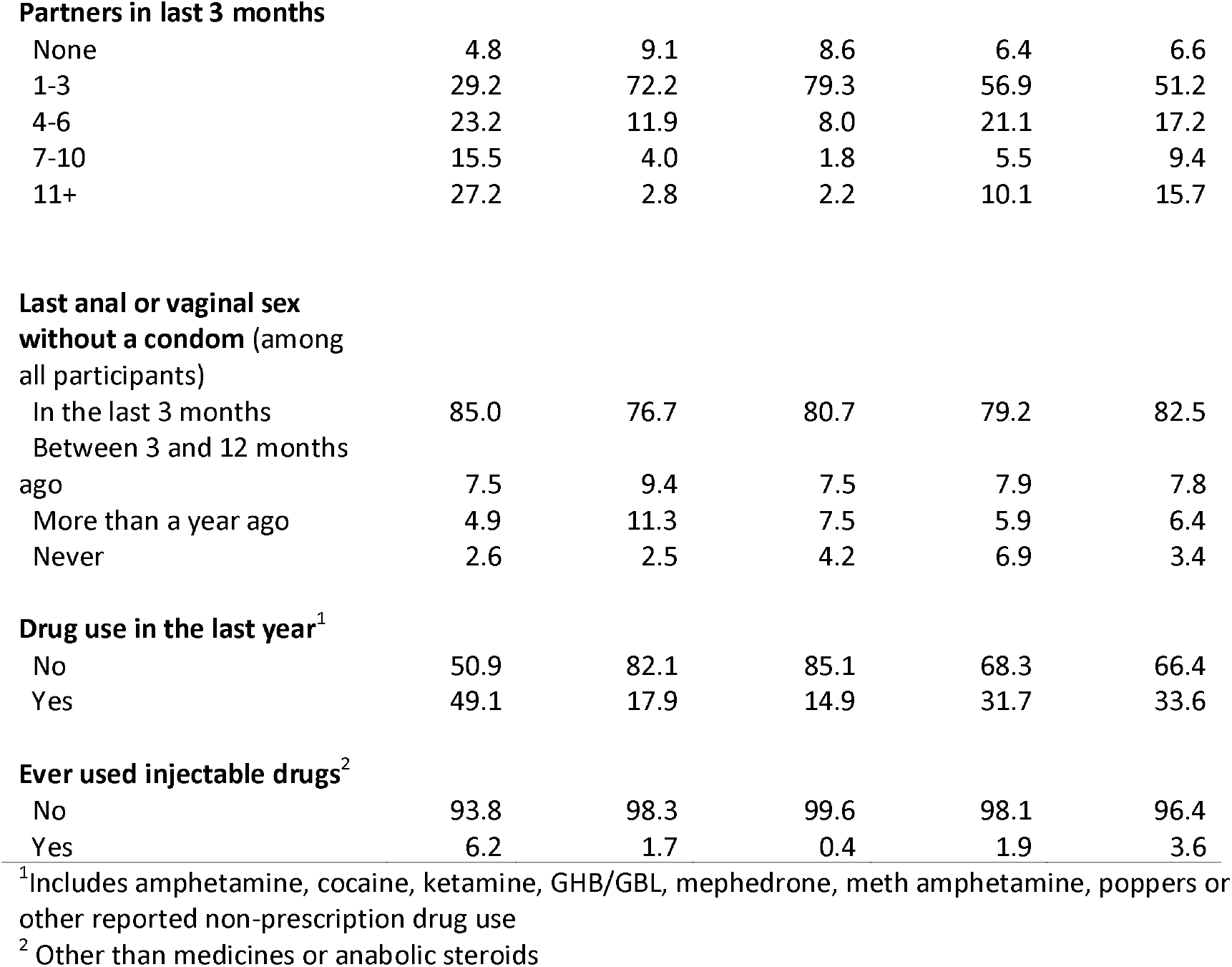
Demographic and behavioural characteristics of the study sample by gender and sexual orientation category.

|  | Gender and sexual orientation category |  |  |  |  |
| --- | --- | --- | --- | --- | --- |
|  | GBMSM<br>(n=866) | MSW<br>(n=201) | Women<br>(n=536) | Other/<br>prefer not | Total<br>(1,723) |
| % |  |  |  |  |  |

|  |  |  |  | to say<br>(n=120) |  |
| --- | --- | --- | --- | --- | --- |
| **Gender identity** |  |  |  |  |  |
| Cisgender | 99.2 | 99.0 | 98.0 | 40.0 | 95.0 |
| Transgender | 0.8 | 1.0 | 1.5 | 1.8 | 1.1 |
| Non-binary/intersex/other | 0.0 | 0.0 | 0.6 | 40.0 | 2.7 |
| Prefer not to say | 0.0 | 0.0 | 0.0 | 18.2 | 1.2 |
| **Age group** |  |  |  |  |  |
| 16-24 | 7.0 | 23.8 | 31.0 | 16.0 | 17.1 |
| 25-34 | 33.2 | 50.6 | 47.3 | 39.6 | 40.0 |
| 35-44 | 30.5 | 15.9 | 16.3 | 27.4 | 24.2 |
| 45-54 | 17.3 | 7.3 | 4.7 | 8.5 | 11.56 |
| 55+ | 12.1 | 2.4 | 0.8 | 8.5 | 7.2 |
| **Ethnicity** |  |  |  |  |  |
| White | 79.8 | 63.2 | 62.7 | 60.8 | 71.2 |
| Black | 3.6 | 18.9 | 12.1 | 10.8 | 8.5 |
| Asian | 7.4 | 11.0 | 11.8 | 5.0 | 9.0 |
| Mixed | 5.9 | 4.0 | 10.8 | 13.3 | 7.7 |
| Other | 3.4 | 3.0 | 2.6 | 10.0 | 3.5 |
| **Born in UK** |  |  |  |  |  |
| No | 45.6 | 34.7 | 38.3 | 43.9 | 42.0 |
| Yes | 54.4 | 65.3 | 61.7 | 56.1 | 58.0 |
| **Clinic Location** |  |  |  |  |  |
| London | 84.2 | 84.6 | 78.5 | 84.2 | 82.5 |
| East of England | 15.8 | 15.4 | 21.5 | 15.8 | 17.5 |
| **Years of education since age 16** |  |  |  |  |  |
| None | 4.7 | 3.5 | 4.9 | 10.0 | 5.0 |
| Up to 2 years | 7.5 | 12.9 | 11.9 | 16.7 | 10.2 |
| 2 years or more | 81.8 | 72.1 | 68.3 | 65.8 | 75.3 |
| Still in full time education | 6.0 | 11.4 | 14.9 | 7.5 | 9.5 |
| **Employed** |  |  |  |  |  |
| No | 15.8 | 19.4 | 22.6 | 30.0 | 19.3 |
| Yes | 84.2 | 80.6 | 77.4 | 70.0 | 80.7 |
| **HIV status** |  |  |  |  |  |
| Never tested | 2.0 | 32.1 | 33.9 | 21.9 | 16.6 |
| Living with HIV | 20.9 | 2.2 | 0.6 | 10.5 | 11.8 |
| HIV negative | 77.1 | 65.8 | 65.6 | 67.5 | 71.6 |
| **STI diagnosis in the last year** |  |  |  |  |  |
| No | 48.4 | 76.2 | 73.8 | 62.4 | 60.3 |
| Yes | 50.0 | 18.8 | 23.8 | 33.0 | 37.3 |
| Not sure | 1.6 | 5.0 | 2.4 | 4.6 | 2.4 |
| **Partners in last 3 months** |  |  |  |  |  |
| None | 4.8 | 9.1 | 8.6 | 6.4 | 6.6 |
| 1-3 | 29.2 | 72.2 | 79.3 | 56.9 | 51.2 |
| 4-6 | 23.2 | 11.9 | 8.0 | 21.1 | 17.2 |
| 7-10 | 15.5 | 4.0 | 1.8 | 5.5 | 9.4 |
| 11+ | 27.2 | 2.8 | 2.2 | 10.1 | 15.7 |
| **Last anal or vaginal sex without a condom (among all participants)** |  |  |  |  |  |
| In the last 3 months | 85.0 | 76.7 | 80.7 | 79.2 | 82.5 |
| Between 3 and 12 months ago | 7.5 | 9.4 | 7.5 | 7.9 | 7.8 |
| More than a year ago | 4.9 | 11.3 | 7.5 | 5.9 | 6.4 |
| Never | 2.6 | 2.5 | 4.2 | 6.9 | 3.4 |
| **Drug use in the last year<sup>1</sup>** |  |  |  |  |  |
| No | 50.9 | 82.1 | 85.1 | 68.3 | 66.4 |
| Yes | 49.1 | 17.9 | 14.9 | 31.7 | 33.6 |
| **Ever used injectable drugs<sup>2</sup>** |  |  |  |  |  |
| No | 93.8 | 98.3 | 99.6 | 98.1 | 96.4 |
| Yes | 6.2 | 1.7 | 0.4 | 1.9 | 3.6 |

<sup>1</sup>Includes amphetamine, cocaine, ketamine, GHB/GBL, mephedrone, meth amphetamine, poppers or other reported non-prescription drug use
<sup>2</sup> Other than medicines or anabolic steroids

The demographic characteristics of all attendees across the study sites during the recruitment period is given in Supplementary Table 1. The study sample was broadly similar to the overall clinic population, however the study sample was less likely to contain Black attendees and more likely to include people living with HIV.

### Awareness and use of HIV PrEP

74% of participants reporting having heard of HIV PrEP (Table 2). This was highest among GBMSM (98%) and lower in MSW (39%) and women (49%). Awareness was highest among people living with HIV (94% vs 72% if negative or unknown status), White participants (78%; Black: 55%; Asian: 66%; Mixed: 66%), those attending SHS in London (77% vs 61%) and participants diagnosed with an STI in the last year (83% vs 71%). Awareness was lower in the 16–24-year-old age group (53%) compared to other age categories (25-34: 73%; 35-44: 80%; 45-54: 86%; 55+: 88%) (not reported in tables).

**Table 2:**
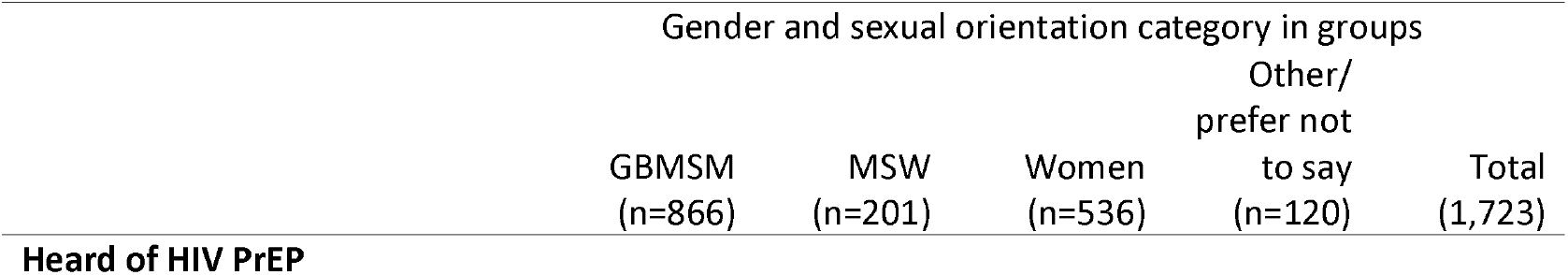
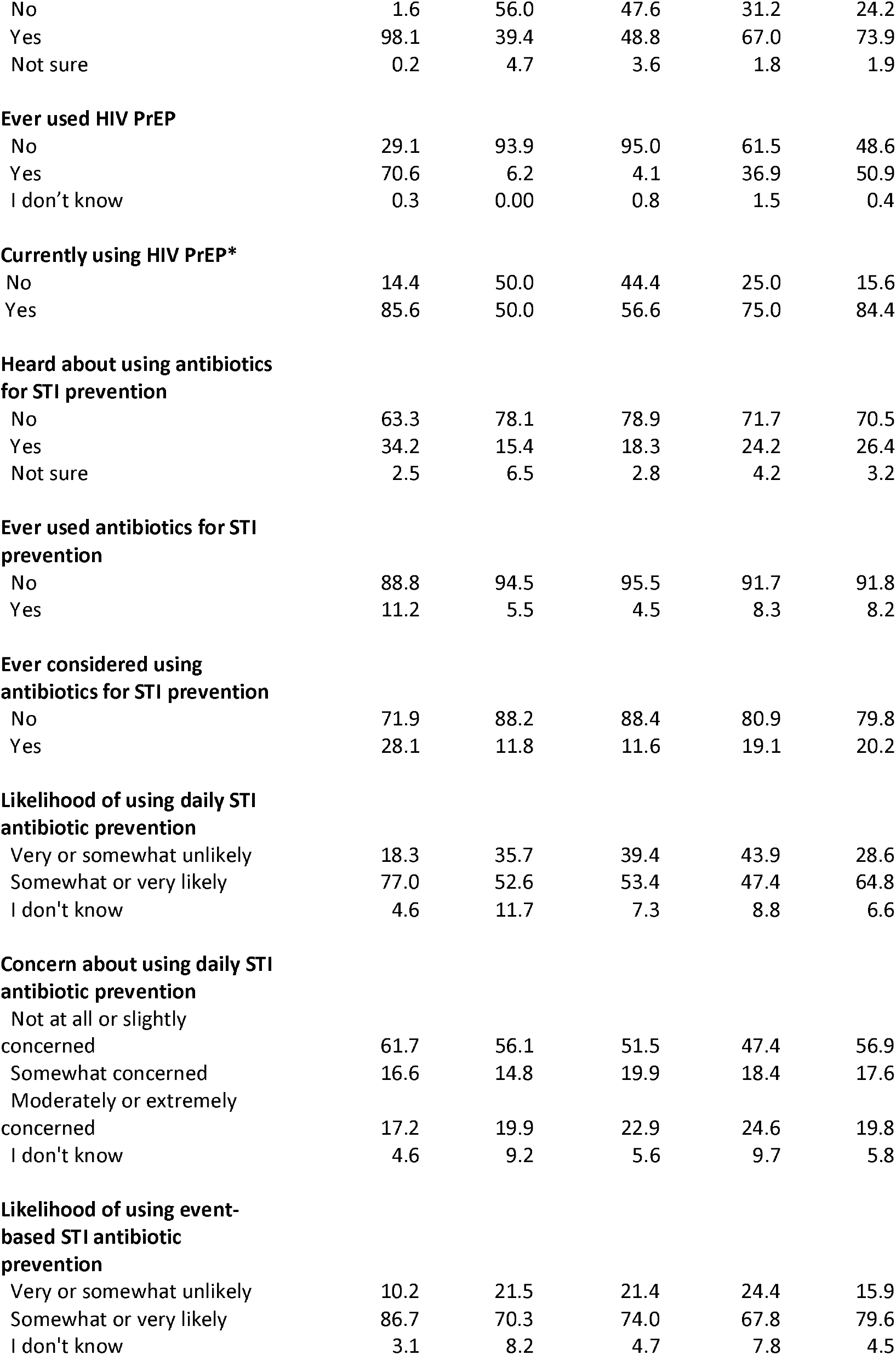
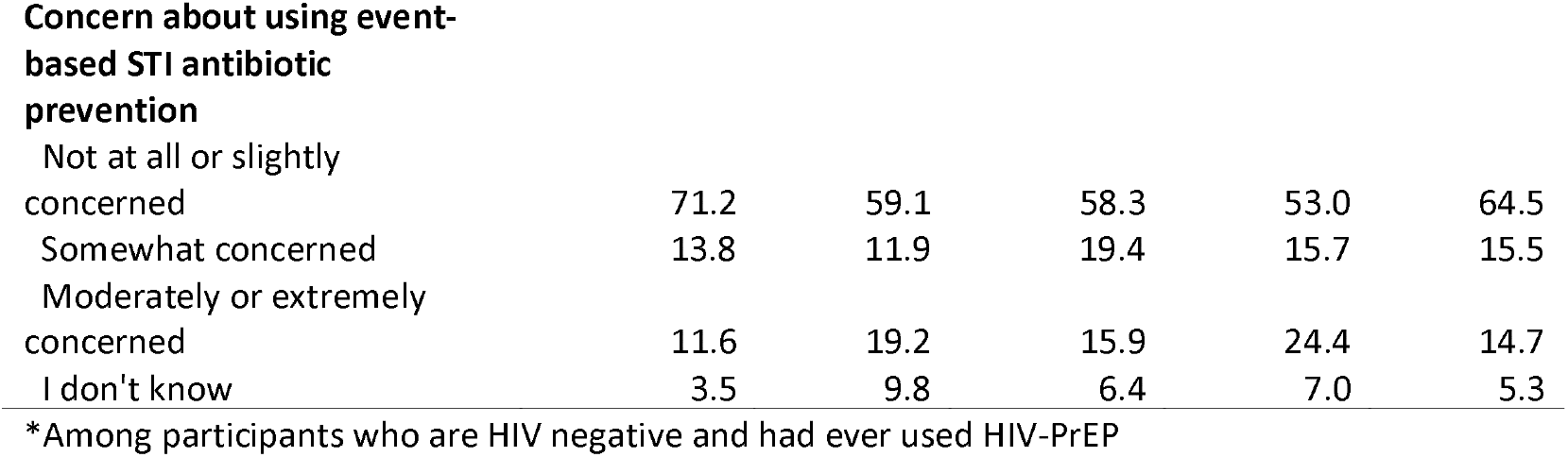
Awareness and use of HIV PrEP and awareness, use, likelihood to use and concerns with antibiotic use for STI prevention by gender and sexual orientation category.

|  | Gender and sexual orientation category in groups |  |  |  | Total<br>(1,723) |
| --- | --- | --- | --- | --- | --- |
|  | GBMSM<br>(n=866) | MSW<br>(n=201) | Women<br>(n=536) | Other/<br>prefer not<br>to say<br>(n=120) |  |
| **Heard of HIV PrEP** |  |  |  |  |  |
| No | 1.6 | 56.0 | 47.6 | 31.2 | 24.2 |
| Yes | 98.1 | 39.4 | 48.8 | 67.0 | 73.9 |
| Not sure | 0.2 | 4.7 | 3.6 | 1.8 | 1.9 |
| **Ever used HIV PrEP** |  |  |  |  |  |
| No | 29.1 | 93.9 | 95.0 | 61.5 | 48.6 |
| Yes | 70.6 | 6.2 | 4.1 | 36.9 | 50.9 |
| I don't know | 0.3 | 0.00 | 0.8 | 1.5 | 0.4 |
| **Currently using HIV PrEP\*** |  |  |  |  |  |
| No | 14.4 | 50.0 | 44.4 | 25.0 | 15.6 |
| Yes | 85.6 | 50.0 | 56.6 | 75.0 | 84.4 |
| **Heard about using antibiotics for STI prevention** |  |  |  |  |  |
| No | 63.3 | 78.1 | 78.9 | 71.7 | 70.5 |
| Yes | 34.2 | 15.4 | 18.3 | 24.2 | 26.4 |
| Not sure | 2.5 | 6.5 | 2.8 | 4.2 | 3.2 |
| **Ever used antibiotics for STI prevention** |  |  |  |  |  |
| No | 88.8 | 94.5 | 95.5 | 91.7 | 91.8 |
| Yes | 11.2 | 5.5 | 4.5 | 8.3 | 8.2 |
| **Ever considered using antibiotics for STI prevention** |  |  |  |  |  |
| No | 71.9 | 88.2 | 88.4 | 80.9 | 79.8 |
| Yes | 28.1 | 11.8 | 11.6 | 19.1 | 20.2 |
| **Likelihood of using daily STI antibiotic prevention** |  |  |  |  |  |
| Very or somewhat unlikely | 18.3 | 35.7 | 39.4 | 43.9 | 28.6 |
| Somewhat or very likely | 77.0 | 52.6 | 53.4 | 47.4 | 64.8 |
| I don't know | 4.6 | 11.7 | 7.3 | 8.8 | 6.6 |
| **Concern about using daily STI antibiotic prevention** |  |  |  |  |  |
| Not at all or slightly concerned | 61.7 | 56.1 | 51.5 | 47.4 | 56.9 |
| Somewhat concerned | 16.6 | 14.8 | 19.9 | 18.4 | 17.6 |
| Moderately or extremely concerned | 17.2 | 19.9 | 22.9 | 24.6 | 19.8 |
| I don't know | 4.6 | 9.2 | 5.6 | 9.7 | 5.8 |
| **Likelihood of using event-based STI antibiotic prevention** |  |  |  |  |  |
| Very or somewhat unlikely | 10.2 | 21.5 | 21.4 | 24.4 | 15.9 |
| Somewhat or very likely | 86.7 | 70.3 | 74.0 | 67.8 | 79.6 |
| I don't know | 3.1 | 8.2 | 4.7 | 7.8 | 4.5 |
| **Concern about using event-based STI antibiotic prevention** |  |  |  |  |  |
| Not at all or slightly concerned | 71.2 | 59.1 | 58.3 | 53.0 | 64.5 |
| Somewhat concerned | 13.8 | 11.9 | 19.4 | 15.7 | 15.5 |
| Moderately or extremely concerned | 11.6 | 19.2 | 15.9 | 24.4 | 14.7 |
| I don't know | 3.5 | 9.8 | 6.4 | 7.0 | 5.3 |

\*Among participants who are HIV negative and had ever used HIV-PrEP

In total, 51% of participants had ever used HIV PrEP, of whom 84% were using it at the time of the survey. Those who had ever used HIV PrEP were more likely to be GBMSM (71% vs 6% of MSW and 4% of women), White (55%; Black: 24%; Asian: 40%; Mixed: 45%), attending SHS in the East of England (59% vs 49%) and diagnosed with an STI in the last year (68% vs 40%). Those in the 16–24-year-old age group were less likely to have ever used HIV PrEP compared to older groups (16-24: 30%; 25-34: 50%; 35-44: 61%; 45-54: 55%; 55+: 44%.

### Awareness and use of antibiotics for STI prophylaxis

Of all participants, 26% had heard of using antibiotics for STI prevention (34% of GBMSM, 15% of MSW and 18% of women) and 8% had ever used antibiotics for STI prevention. Use was highest among GBMSM (11%) and lower among MSW (5%) and women (4%). Awareness and use were highest among people living with HIV (35%;15%), those attending SHS in London (28%; 9%) and participants diagnosed with an STI in the last year (34%; 12%). Use was lowest in 16–24-year-olds (6%) and highest in 25-44 year-olds (12%). Despite fluctuations, awareness and use were largely consistent across the period of recruitment (Supplement 1).

A small number of participants (n=31) may have misunderstood the meaning of STI PEP/PrEP because some participants gave responses consistent with antibiotic treatment for STIs. Accounting for this in a sensitivity analysis, 6% of the study sample had ever used antibiotics for STI prevention (9% of GBMSM, 5% of MSW and 2% of women). Those who had potentially misunderstood the question were more likely to be GBMSM (52%) or WSM (42%) compared to MSW (3%).

### Details of antibiotic use to prevent STIs

Of the participants who had previously used antibiotics for STI prophylaxis (n=142), 39% had discussed their use with a healthcare professional. 61% took doxycycline, 13% used azithromycin, 12% used amoxicillin and 5% used metronidazole. 39% bought the antibiotics online and 55% had them prescribed for this or another purpose. Three participants reported being given them by a partner or a friend. 45% first took antibiotics to prevent STIs more than a year before the survey date, and 57% had used antibiotics in this way in the three months before the survey. 47% took two antibiotics pills in the 24 hours after sex (STI PEP), 28% took daily doxycycline (STI PrEP), and 24% adopted a different dosing schedule. Of the 23 participants who gave details of their alternative dosing regimen, one took 200mg doxycycline within 72 hours of sex (i.e. doxyPEP), whereas the remainder adopted unstudied regimens. Common dosing practices included taking doxycycline for between 3 and 10 days after exposure (n=10) or using a lower dose of doxycycline than recommended (n=7). 76% said they were trying to prevent chlamydia, 55% gonorrhoea and 42% syphilis. 9% said they were trying to prevent LGV, 6% herpes, 6% shigella and 2% hepatitis.

When asked why they decided to take antibiotic prophylaxis (n=142), 21% said they decided to take it after risky behaviour, and 17% said it was to prevent STI infections. 13% took it based on the scientific evidence available for its efficacy, 9% referred to STI treatment, 5% were recommended by a healthcare provider, 7% heard about it from friends and 6% took it because they were getting frequent STIs. 5% took it based on recommendations in other countries and 5% took it pre-emptively after a partner tested positive for an STI.

### Likelihood to use and concerns with antibiotic prophylaxis use

Of participants who had not used STI prophylaxis (regardless of whether they had previously heard of STI PEP/PrEP), 20% had considered doing so (28% of GBMSM, 12% of MSW and 12% of women). Consideration of use was highest among people living with HIV (30%) and those who were diagnosed with an STI in the last year (30%). Participants were more likely to use event-based dosing and had fewer concerns with this dosage method compared to daily dosing (Table 2). GBMSM expressed a higher likelihood to use and fewer concerns with antibiotic prophylaxis compared to MSW or women.

### Regression modelling results for use of STI PEP/PrEP

The results of the demographic regression analysis provided no evidence of an association between prophylactic use of antibiotics to prevent STIs and ethnicity, place of birth, level of education and employment status (Table 3). Both the bivariate and multivariable models showed an association between antibiotic use and age: 35-44 year olds had higher odds of use than 25-34 year olds (aOR: 1.81 (95% CI: 1.12-2.93)). Compared to GBMSM, MSW (0.56 (0.27-1.14)) and women (0.42 (0.25-0.72)) had a lower odds of using antibiotic prophylaxis in the adjusted model. In the unadjusted model, participants in the East of England had a lower odds of use compared to those attending a SHS in London (0.53 (0.31-0.92)), but this association was attenuated in the final model (0.71 (0.39-1.29)).

**Table 3:** Unadjusted and adjusted logistic regression models for use of antibiotics to prevent STIs among all survey participants.

| Variable | Category | Unadjusted model<br>OR (95% CI) | Adjusted model<br>OR (95% CI) (n=1,483) |
| --- | --- | --- | --- |
| **Age group** (n = 1,532) | 16-24 | 0.92 (0.5-1.7) | 0.97 (0.49-1.95) |
|  | 25-34 | 1 | 1 |
|  | 35-44 | 1.99 (1.26-3.14) | 1.81 (1.12-2.93) |
|  | 45-54 | 1.71 (0.95-3.08) | 1.26 (0.67-2.4) |
|  | 55+ | 1.35 (0.63-2.87) | 1.02 (0.46-2.29) |
| **Gender and sexual orientation** (n=1723) | GBMSM | 1 | 1 |
|  | MSW | 0.46 (0.24-0.87) | 0.56 (0.27-1.14) |
|  | Women | 0.37 (0.23-0.59) | 0.42 (0.25-0.72) |
|  | Other/ prefer not to say | 0.72 (0.36-1.42) | 0.69 (0.31-1.5) |
| **Ethnicity** (n=1721) | White | 1 | 1 |
|  | Black | 1.18 (0.66-2.13) | 1.72 (0.88-3.37) |
|  | Asian | 0.61 (0.29-1.28) | 0.8 (0.37-1.74) |
|  | Mixed | 1.54 (0.88-2.7) | 1.52 (0.79-2.94) |
|  | Other/ prefer not to say | 0.79 (0.28-2.22) | 0.89 (0.31-2.61) |
| **Born in the UK** (n=1,647) | No | 1.1 (0.77-1.57) | 0.86 (0.57-1.29) |
|  | Yes | 1 | 1 |
| **SHS location** (n=1723) | London | 1 | 1 |
|  | East of England | 0.53 (0.31-0.92) | 0.71 (0.39-1.29) |
| **Education** (n=1723) | Up to two years | 0.78 (0.46-1.3) | 0.92 (0.5-1.68) |
|  | More than two years | 1 | 1 |
|  | In education | 0.75 (0.4-1.43) | 1.23 (0.57-2.64) |
| **Employed** (n=1,718) | Yes | 1.03 (0.67-1.58) | 1.22 (0.73-2.03) |
|  | No | 1 | 1 |

In the behavioural regression analysis among GBMSM, reporting an STI diagnosis in the last year (1.82 (1.16-2.85)), eleven or more partners in the last three months (1.79 (1.13-2.82)), sexualised drug use in the last year (1.63 (1.06-2.5)) and living with HIV (2.29 (1.21-4.35)) were independently associated with antibiotic use to prevent STIs (Table 4). In the fully adjusted model,reporting at least two markers of higher-risk behaviour (an STI in the last year, more than 11 partners in the last 3 months, or sexualised drug use or injectable drug use) was associated with antibiotic prophylaxis use (2.11 (1.29-3.48)), but living with HIV was not (1.79 (0.80-4.01)). Although those taking HIV PrEP were more likely to report using antibiotics in this way than those who were not taking HIV PrEP, the 95% confidence interval crossed the null value (1.18 (0.61-2.28)).

**Table 4:** Unadjusted and adjusted logistic regression models for use of antibiotics to prevent STIs among GBMSM.

| Variable | Category | Bivariate model<br>OR (95% CI) | Multivariable<br>model<br>OR (95% CI)*<br>(n=747) |
| --- | --- | --- | --- |
| **STI diagnosis in the last year** (n=830) | Yes | 1.82 (1.16-2.85) |  |
|  | No | 1 |  |
| **Number of partners in the last 3 months** (n=831) | None | 0.24 (0.03-1.80) |  |
|  | One to ten | 1 |  |
|  | Eleven or more | 1.79 (1.14-2.82) |  |
| **Sexualised drug use in the last year** (n=866) | Yes | 1.63 (1.06-2.5) |  |
|  | No | 1 |  |
| **Ever injected drugs** (n=825) | Yes | 1.81 (0.85-3.85) |  |
|  | No | 1 |  |
| **HIV/PrEP status** (n=842) | Negative/unknown, not taking PrEP | 1 | 1 |
|  | Negative/unknown, taking PrEP | 1.61 (0.9-2.86) | 1.18 (0.61-2.28) |
|  | Living with HIV | 2.29 (1.21-4.35) | 1.79 (0.80-4.01) |
| **Higher risk (composite)** (n=866) | Yes | 2.1 (1.37-3.21) | 2.11 (1.29-3.48) |
|  | No | 1 | 1 |

\*Controlled for age group, ethnicity and clinic location

## Discussion

This study used survey data collected from SHS attendees in London and the East of England to investigate awareness and use of biomedical HIV and STI prophylaxis. We found that although awareness of HIV-PrEP was high among GBMSM, it was considerably lower in MSW and women. 34% of GBMSM had heard of STI PEP/PrEP (compared to 15% of MSW and 18% of women). 11% of GBMSM had used antibiotics for STI prophylaxis (compared to 5% of MSW and 4% of women). Participants reported a high likelihood to use STI PEP/PrEP in the future, with a preference for event-based dosing. STI PEP/PrEP use among GBMSM was associated with reporting higher-risk sexual behaviours (a recent STI diagnosis, larger partner numbers in the recent past and/or use of sexualised or injectable drugs).

### Strengths and Limitations

This study has several strengths. Much of the existing evidence on use of STI PEP/PrEP comes from GBMSM populations (3-5), but this study included and therefore is able to demonstrate that heterosexual men and women are also using STI PEP/PrEP and report high interest in using it in future. The study population of SHS attendees was appropriate for the outcome of interest as it was more likely to include people who engage in higher-risk sexual behaviour than a sample of the general population(22). The survey contributes to the growing evidence base around STI PEP/PrEP use (13-15, 17) and extends it by reporting in detail for different population groups on current use of antibiotics for STI prevention (including dosing schedule, source of antibiotics and reasons for taking them).

The results are subject to some limitations. Small numbers of MSW and women reported taking STI PEP/PrEP limit the conclusions we can reach about the factors associated with antibiotic prophylaxis and current patterns of use among these groups. The estimate of STI PEP/PrEP use may have been affected by participant misunderstanding of the question. A sensitivity analysis was conducted to understand the extent of this potential misclassification, which showed little impact on the overall findings. This suggests that additional pilot studies and cognitive testing may be warranted to assess the validity of survey instruments on this topic, and to ensure that participants understand the definition of STI PEP/PrEP presented in the questionnaire. Small numbers of transgender and gender-diverse participants limited specific analysis of these populations.

The study findings may have been affected participation bias (i.e. those who were already aware of or interested in STI PEP/PrEP may have been more likely to take part) or a priming effect, by which taking part in the survey made them aware of STI PEP/PrEP. Finally, this was a cross-sectional study, which means we cannot determine the temporal relationship between risk behaviour and use of antibiotic STI prophylaxis.

### Comparison with existing evidence

The study found low awareness and use of HIV PrEP among MSW (39%), women (49%), participants of Black ethnicity (55%) and those aged 16-24 (53%). Despite methodological differences, these results are in line with a study which reported low awareness of HIV PrEP among heterosexual men and women in the United States (23) and a systematic review on HIV PrEP provision in Europe which observed low levels of awareness among women in six studies (24). Lower use of PrEP has also been documented among Black populations (25, 26) and younger age groups (23, 26). These groups may have low HIV PrEP awareness because clinicians do not provide information during SHS visits due to a perception that HIV PrEP is not suitable for certain demographics, or because of wider inequalities which prevent some individuals from accessing SHS. Our results also suggest that MSW and women have (or at least report) fewer markers of higher-risk activity than GBMSM, and so are likely to have (or be perceived as having) a lower need for HIV-PrEP, which could mean healthcare providers are less likely to discuss HIV-PrEP with them, therefore explaining why PrEP awareness is lower in these groups.

The results also suggest that participants visiting SHS London are less likely to take HIV PrEP than those who attended an SHS in the East of England. This may be explained by the higher proportion of White participants in the East of England, who are more likely than other ethnicities to access HIV PrEP.

Estimates of STI PEP/PrEP use among GBMSM are broadly consistent with existing literature. Surveys of GBMSM and HIV PrEP users in the United States, UK, European Union and Australia between 2018-2022 report that 2-10% of those surveyed had used antibiotics for STI prevention (27). In 2023, surveys of GBMSM and users of sexual networking apps in Germany and the United States reported 23% - 21% of participants had used STI PEP/PrEP (18, 19). Our results did not show a large increase in use in 2023, perhaps due to differing methodologies or divergent patterns of STI PEP/PrEP use in the UK compared to other parts of the world. It is also possible that media coverage of resistance gonorrhoea has been more comprehensive in the UK than in the United States or Germany, which also could have influenced use (28). The use of antibiotics other than doxycycline is consistent with surveys from other settings (13, 15). This suggests that a clinician-led model for STI PEP/PrEP may be needed to reduce the risk of antibiotic resistance through self-sourcing, although community education on antimicrobial resistance may also be effective.

The regression results confirm that GBMSM are more likely to report STI PEP/PrEP use than other population groups, and that GBMSM who report engaging in higher risk activities are more likely to use antibiotics in this way. HIV negative GBMSM using HIV-PrEP were not more likely to do so than HIV negative participants not using HIV-PrEP (29). This finding may be due to the relatively small sample size of HIV-PrEP users, meaning the analysis was under-powered to demonstrate an effect.

### Implications

The study results have implications for SHS in England. Consideration could be given to how to more effectively inform and encourage the use of HIV-PrEP where appropriate among MSW, women, younger age groups, Black populations and other marginalised groups about HIV-PrEP, bearing in mind that the need for PrEP may vary considerably across these groups. There is also no widely agreed markers of HIV-PrEP need in women, which makes the level of need difficult to quantify. Most STI PEP/PrEP users are taking antibiotics in accordance with that used in clinical trials, but some report using non-recommended regimens. However although some participants may have erroneously reported regimens used for treatment rather than prophylaxis. Some participants report using an antibiotic other than doxycycline, following a dosing regimen other than daily or event-based. A large proportion report that they are using antibiotics to prevent gonorrhoea. Implementation of STI PEP/PrEP should contain messaging to address misconceptions about which antibiotics can be used for this purpose, the appropriate dosing schedule, and to inform potential users of the risk of antimicrobial resistance.

Acceptability of STI PEP/PrEP was high across all participant groups, meaning that any future research and implementation should consider the needs and preferences of heterosexual men and women, not just GBMSM populations. We recommend further research among population groups at higher risk other than GBMSM given the limitations of this survey in understanding these populations.

## Conclusion

This survey of attendees of SHS in London and East of England showed that awareness of HIV PrEP was high among GBMSM, but considerably lower in MSW and women. A minority of participants had used STI PEP/PrEP, but interest in doing so in the future was high among all groups. The results suggest that more consideration should be given to raising awareness of and identifying a need for HIV-PrEP among non-GBMSM groups. They also have implications for future STI PEP/PrEP introduction: messaging should encourage evidence-based use of antibiotics for STI prevention and guidelines should account for the needs of MSW and cisgender women.

## Data Availability

Data are available upon reasonable request.

## Acknowledgements

We would like to acknowledge the contribution of Roeann Osman, who provided supplementary analysis from the GUMCAD dataset.

We acknowledge members of the National Institute for Health and Care Research Health Protection Research Unit (NIHR HPRU) in Blood Borne and Sexually Transmitted Infections (BBSTI) Steering Committee: Professor Caroline Sabin (HPRU Director), Dr John Saunders (UKHSA Lead), Professor Catherine Mercer, Professor Gwenda Hughes, Dr Hamish Mohammed, Professor Greta Rait, Dr Ruth Simmons, Professor William Rosenberg, Dr Tamyo Mbisa, Professor Rosalind Raine, Dr Sema Mandal, Dr Rosamund Yu, Dr Samreen Ijaz, Dr Fabiana Lorencatto, Dr Rachel Hunter, Dr Kirsty Foster and Dr Mamooma Tahir.

