## Supplementary Table 1 for "Prevalence and factors associated with use of prophylaxis for HIV and sexually transmitted infections among sexual health service attendees in England: findings from a cross-sectional observational study"

**Supplementary Table 1: The demographic characteristics of all attendees at the study sites during the recruitment period**

|  | **Gender and sexual orientation category** | | | | |
| --- | --- | --- | --- | --- | --- |
| % | GBMSM  (n=73,192) | MSW  (n=49,225) | Women  (n=145,265) | Not known  (n=51,344) | Total  (319,026) |
| **Gender identity** | | | | | |
| Cisgender | 99.56 | 99.05 | 98.81 | 96.34 | 98.62 |
| Transgender | 0.44 | 0.95 | 1.19 | 1.09 | 0.97 |
| Gender diverse | 0.00 | 0.00 | 0.00 | 2.57 | 0.41 |
| **Age group** | | | | | |
| 16-24 | 6.22 | 19.59 | 28.05 | 22.14 | 20.79 |
| 25-34 | 25.85 | 37.36 | 39.41 | 36.87 | 35.57 |
| 35-44 | 24.73 | 20.01 | 19.08 | 21.23 | 20.87 |
| 45-54 | 21.97 | 11.50 | 9.27 | 11.56 | 12.89 |
| 55+ | 21.22 | 11.54 | 4.19 | 8.20 | 9.88 |
| **Ethnicity** | | | | | |
| White | 67.59 | 49.55 | 54.93 | 57.32 | 57.39 |
| Black | 4.82 | 18.93 | 14.94 | 6.21 | 11.83 |
| Asian | 7.91 | 7.27 | 7.25 | 6.66 | 7.31 |
| Mixed | 5.65 | 5.72 | 6.91 | 4.74 | 6.09 |
| Other | 2.72 | 1.90 | 1.78 | 1.50 | 1.97 |
| Unknown | 11.31 | 16.63 | 14.20 | 23.56 | 15.42 |
| **Born in UK** | | | | | |
| No | 40.09 | 30.45 | 30.72 | 23.32 | 31.64 |
| Yes | 43.06 | 54.43 | 57.04 | 53.51 | 52.86 |
| Unknown | 16.85 | 15.11 | 12.24 | 23.17 | 15.50 |
| **Clinic Location** | | | | | |
| London | 89.17 | 75.72 | 69.88 | 75.48 | 76.11 |
| East of England | 10.83 | 24.28 | 30.12 | 24.52 | 23.89 |
| **HIV status** | | | | | |
| Living with HIV | 4.52 | 1.54 | 0.63 | 1.28 | 1.77 |
| HIV negative | 95.48 | 98.46 | 99.37 | 98.72 | 98.23 |
| **STI diagnosis in the last year** | | | | | |
| No | 60.76 | 66.98 | 65.19 | 58.90 | 62.65 |
| Yes | 39.24 | 33.02 | 34.81 | 41.10 | 37.35 |

**Notes:**

- Data has been deduplicated at the patient level (i.e., each count corresponds to a one patient)
- The values for gender identity, age group, ethnicity, and country of birth were derived from the coded value at the patients first appointment within the study recruitment period for that clinic.
- HIV prevalence was determined based on whether a “new HIV diagnosis” code ever appeared in a patients record before/ last day of study recruitment at the clinic. This may overestimate prevalence, as it may include diagnoses that occurred after patient survey completion date but before the end of the study.
- Previous STI diagnoses in the last year include chlamydia, gonorrhoea, syphilis, herpes (first episode), LGV and hepatitis C.
- Homerton Healthcare clinics (Ivy Centre, 80 Leadenhall Street, John Scott Health Centre and Clifden Centre (Homerton Hospital)) used the earliest and latest study recruitment date for combined sites.
- Wisbech (Vancouver House) and King’s Lynn used the earliest and latest study recruitment date for combined sites.
- Clinics from Milton Keynes (Milton Keynes iCaSH, iCaSH Oak Tree, iCaSH North Cambridgeshire) all submit GUMCAD data using the same clinic code, therefore the earliest and latest study recruitment dates were used for combined sites.
